# How Does Psilocybin Affect Daily Life Experience and Behavior? Protocol For a Multimethod Ambulatory Assessment Study Within a Clinical Trial of Psilocybin for Depression

**DOI:** 10.64898/2026.11.10.26367628

**Authors:** Andreea Sutu, Deanna Kaplan, Charles L. Raison, Christina J. Sauder, Santiago Jose Arconada Alvarez, Nicole S. Pozzo, Zainab Salako, Zhuo Job Chen, Roman Palistky

## Abstract

**Introduction:** Although psilocybin has shown promise for treating depression, responses vary considerably, with some individuals experiencing limiting improvement and even adverse effects. Research has largely relied on assessments conducted at discrete time points, providing limited insight into how post-psilocybin change unfolds in everyday life. The Dial-In study aims to characterize daily-life processes associated with differential responses to psilocybin, including factors associated with improvement or worsening following challenging dosing experiences.

**Methods and Analysis:** Dial-In is a prospective, observational, multi-method longitudinal study embedded within a clinical trial administering 25 mg of psilocybin to up to 141 adults with major depressive disorder. Ambulatory assessments combine ecological momentary assessment (EMA) and voice diaries for 14 days pre- and post-dosing, and passive audio sampling via the Electronically Activated Recorder (EAR) for three four-day periods to characterize everyday affect, behavior, reflections, and social interactions. Outcomes include depressive symptoms, psychological well-being, and psychedelic-related adverse effects assessed through eight weeks post-dose. Analyses will examine whether affective sudden gains and losses, social support, interpersonal conflict, conversational focus, reflection and rigidity predict differential outcomes. Additional analyses will test whether these factors moderate associations between challenging dosing experiences and clinical outcomes.

**Ethics and Dissemination:** The project was approved by WCG Institutional Review Board, with Emory University relying on WCG as the reviewing board. All participants provide written informed consent. Findings will be disseminated through peer-reviewed publications and scientific presentations.

**Trial Registration:** ClinicalTrials.gov, NCT06512194 (parent trial), OSF (https://osf.io/nx5ky/overview)

**Article Summary:** Strengths and Limitations of this Study

- Strength: Measurement of observed behavior, beyond self-report.
- Strength: The integration of ecological momentary assessment, open-ended voice diaries, and naturalistic behavioral observations provides complementary perspectives on post-acute change.
- Strength: The prospective, longitudinal design enables the examination of within-person affective and behavioral changes surrounding psilocybin administration, including sudden gains and losses that may be obscured by discrete time-point assessments.
- Strength: By examining depressive symptoms, psychological well-being, and adverse effects, the study captures multiple dimensions of treatment response.
- Limitation: The observational design precludes causal inference regarding the direction of relationships between everyday processes and clinical outcomes.

---

Psilocybin has been shown to reduce symptoms of depression in multiple studies [1–6], yet responses to psilocybin vary considerably across individuals [2, 4, 7]. Many individuals experience sustained psychological benefits extending beyond symptom reduction [7–10], whereas others experience limited improvement [2, 4] or, at times, worsening across emotional, interpersonal, or spiritual domains [7, 11, 12]. However, a notable absence of ambulatory assessment has made it difficult to ground such research in experiences and behaviors that occur outside the lab, where many of the enduring impacts of psilocybin are expected to unfold.

Ambulatory assessment encompasses methods that collect data in the context of participants’ everyday environment outside of the laboratory, including experience sampling or ecological momentary assessment (EMA), naturalistic observation, and passive behavioral or physiological measurement [13, 14]. These methods can capture data from participants’ behaviors and environments, as well as fine-grained assessments of emotional and cognitive processes, in real or near-real time as they unfold in daily life. The collection of multiple daily assessments also generates intensive longitudinal data streams suitable for determining temporal dynamics of change [14, 15]. The collection of ambulatory data is nascent in psychedelic research, with only one published study to our knowledge having used EMA for full-dose psychedelics [16] (although see [17] for an EMA study of microdosing, and also [18], and clinical trial registrations NCT06355414; NCT06442423 for current protocols that use EMA). Nevertheless, published clinical psilocybin research continues to rely primarily on laboratory assessments or occasional at-home self-report surveys [2–4, 6]. Naturalistic observational studies likewise often rely on waves of surveys delivered either retrospectively [19] or prospectively [7, 20]. Though useful in many ways, these yield limited information about daily and momentary experiences and behaviors, leaving the temporal and contextual dynamics comparatively understudied in psychedelics.

Nonetheless, processes and outcomes captured through ambulatory assessment are highly germane to psychedelics research. These include the assessment of patterns in moment-to-moment emotional dynamics, and changes in those patterns; changes in perspective, wellbeing, and phenomenology, which might unfold either gradually or suddenly; the contributions of individuals’ daily environments and interactions to therapeutic change; and observed behavior change. Such data may help characterize the dynamics of change after psilocybin, identify daily-life factors associated with differential response, and detect early indicators of improvement or deterioration. This protocol paper describes the Dial-In Study, which incorporates ambulatory assessment into an existing clinical trial of psilocybin for major depression (OPTIMIZE; ClinicalTrials.gov NCT06512194) to generate novel ambulatory data on daily-life processes surrounding psilocybin treatment and identify predictors of differential response. The Dial-In study will use multi-method prospective longitudinal assessment to generate fine-grained time-series data across four weeks of the clinical trial. The study integrates three complementary modalities of daily-life assessment: intensive longitudinal self-report via EMA, repeated daily open-ended voice diaries, and naturalistic behavioral observation by means of recorded ambient audio. Together, these methods allow us to characterize participants’ affective dynamics, observed behaviors and social interactions, and participants’ reflections of their experiences, while also examining whether these processes are associated with differential response to psilocybin.

## Known predictors of differential response to psilocybin

Growing evidence has identified several correlates of differential response to psilocybin, spanning features of the acute psychedelic experience, baseline psychological and neurobiological characteristics, and aspects of the therapeutic context [21–23]. Subjective positive dosing experiences have been associated with greater benefits, while challenging experiences have been linked with less benefit and even deterioration after psilocybin, albeit inconsistently [7, 20, 21, 24–26]. Importantly, challenging dosing experiences are not uniformly associated with poor outcomes: in a large retrospective survey, most individuals describing their most difficult psilocybin experience reported eventually benefiting from it [19]. More recent work similarly suggests that the outcomes associated with challenging experiences may depend on how those experiences are processed and supported [27–30], underscoring the importance of understanding when such experiences lead to growth versus worsening. Beyond the dosing experience itself, individuals’ daily environments and social contexts may also shape how psilocybin experiences are integrated over time. Although social and interpersonal contexts have been linked to longer-term psychedelic outcomes [20, 31–33], these factors have rarely been examined prospectively and at a fine-grained temporal level. Consequently, little is known about how individual’s daily environments, experiences, and behaviors are related to post-acute trajectories as they unfold in everyday life [34, 35].

## Contribution of Ambulatory Assessment and Dynamic Systems Modeling

The Dial-In Study uses a *multi-method*, *ambulatory*, *intensive longitudinal* assessment approach. *Multi-method assessment* includes subjective psychological assessment using EMA, daily voice diaries, and passive audio acquisition using the Electronically Activated Recorder (EAR). *Ambulatory assessment* involves tracking experiences that unfold in participants’ everyday environments [13]. Because improvement or deterioration take place over time, and within the context of daily life, ambulatory assessment methods are well suited to capturing clinically relevant phenomena outside of the lab. *Intensive longitudinal* assessment involves frequent repeated measurements at relatively short intervals, and at a sufficiently granular time scale, to enable the characterization of processes that unfold dynamically over time [36, 37]. Using intensive longitudinal methods, psychotherapy research has found that improvement and worsening is often a discontinuous process, comprising sudden gains and losses, which are predictive of the ultimate trajectory of change [38, 39]. This may be especially true of psilocybin, which can produce marked and enduring shifts in subjective experience and perspective [40–42]. Because clinical change following psilocybin can emerge rapidly in the post-dosing window, sudden gains and losses may be especially likely and clinically meaningful for psilocybin assisted therapies [6, 43, 44]. Instantiated in day-to-day patterns of emotion, cognition, and behavior, sudden gains and losses are potential early indicators of subsequent clinical change and differential response.

Dynamic complexity theory offers a potentially useful framework for conceptualizing and evaluating the contribution of momentary dynamics to ultimate therapeutic change [45–47] and identifying early warning signs of differential response. From a complex dynamic systems perspective, psychological functioning may be conceptualized as a persistent system of interacting states in continuous exchange with the environment [48]. Psilocybin may disrupt individuals’ habitual response patterns, creating a period of instability during which the system becomes more susceptible to reorganization [46]. Such reorganization may allow new and adaptive patterns of functioning to emerge but may also introduce vulnerability for maladaptive changes. Intensive longitudinal data collected in this study are suitable to complex dynamic systems modeling and thus offer an opportunity to study psilocybin’s contributions to shifts in individuals’ dynamic systems as part of therapeutic change.

## Study Objectives

This study has two complementary objectives. The first is to generate novel data comprising observational metrics of behavior (via passively sensed audio), affective dynamics (via EMA), and longitudinal qualitative data (voice diary) within the context of a Phase II psilocybin trial. This makes possible the second aim, which consists of planned analyses that will evaluate determinants of differential response to psilocybin. For the planned analyses, differential responses to psilocybin are operationalized as differences in depression reduction, wellbeing enhancement, and adverse effects after the psilocybin intervention. Planned analyses will also examine *determinants* of differential response after challenging dosing experiences, specifically moderators of the relationship between challenging experiences and these outcomes. First, we will examine sudden gains and losses in the time series as independent predictors of differential response, with the expectation that sudden gains will precede better responses and sudden losses will precede poorer responses (H1). Second, we will evaluate ecologically assessed predictors of differential response from participants’ observed environments and behaviors, assessed via passive monitoring with the Electronically Activated Recorder (EAR), and naturalistic speech collected via daily voice diaries. Using composite indices derived from these methods, we expect more support, less conflict, and a shift in conversational focus from self to others (EAR; H2a), and greater personal reflection and lower rigidity (voice diary; H2b), to be associated with better outcomes. We expect to replicate previously observed associations between challenging experiences and poorer responses (H3a), but that the risk and protective variables identified in H1-2 will moderate this relationship, with protective factors exerting a buffering role and risk factors strengthening the relationship (H3b). Finally, the resulting longitudinal data will support exploratory analyses of additional temporal and dynamic predictors of differential treatment response using dynamic systems and other computational approaches.

## Methods

The Dial-In study is embedded within a larger clinical trial administering a single 25 mg dose of psilocybin to adults meeting criteria for Major Depressive Disorder. In the parent trial all participants receive 2 preparation sessions with trained therapists, a single dosing session, and 3 integration sessions, comprising 8 hours of interpersonal support. After psilocybin dosing, participants are randomized to receive either transcutaneous auricular vagus nerve stimulation (taVNS), sham-taVNS, or no taVNS. See Supplement A, “Parent Study Design and Procedure” for further detail on the parent study’s design. Methods for the Dial-In Study are described below. The parent protocol and informed consent form were approved by WCG IRB, with Emory University relying on WCG as the reviewing IRB under an institutional reliance agreement. All participants provided written informed consent (ICF) for OPTIMIZE. A separate consent form was not used for Dial-In as its procedures were summarized in the parent ICF. Emory University oversees the collection and processing of the EMA, voice-diary, and EAR data. Identifiable audio data are transferred to and stored on a secure SharePoint folder, with access restricted to authorized study personnel.

### Participants

OPTIMIZE plans to enroll up to 141 adults aged 18–70. Participants must meet the DSM-5 criteria for Major Depressive Disorder (MDD) with a current major depressive episode of at least 60 days’ duration and clinically significant depressive symptoms at screening (as indexed by a MADRS score of ≥ 28). Additional inclusion criteria include medical eligibility for psilocybin administration, willingness to comply with study procedures, and ability to complete assessments in either English or Spanish. The study began enrolling participants on 05/13/2025 and will continue enrolling participants through 05/2029. At the time of submission, the study has enrolled 41 subjects and the recruitment is ongoing.

Exclusion criteria include a history of psychotic or bipolar disorders; current or recent (within 12 months of screening) anorexia nervosa, bulimia nervosa, or antisocial personality disorder; and current or recent obsessive-compulsive disorder, posttraumatic stress disorder, panic disorder, agoraphobia, or binge eating disorder when judged to be primary relative to MDD. Additional exclusion criteria include active suicidal intent or a suicide attempt within the previous 6 months, current substance use disorder or moderate-to-severe alcohol use disorder, regular tobacco use that would preclude the required abstinence period, recent psychedelic use, clinically significant medical contraindications, pregnancy or breastfeeding, and inability or unwillingness to discontinue prohibited psychotropic medications. Participants judged unsuitable for study participation for other reasons at the discretion of the investigators are also excluded. All participants recruited for the parent-study will also follow the Dial-In Study procedures and assessment.

### Study Design & Procedures

The Dial-In study is an observational study embedded within the OPTIMIZE trial and uses four types of data: self-report EMA surveys, daily voice diaries, and passively sensed audio data collected via the EAR, as well as standardized clinical assessments collected under the parent-trial protocol, comprising depression, wellbeing, and adverse effects.

The assessment schedule for this study is represented in Figure 1. For consistency, assessment time points are referred to throughout as Baseline 1, Baseline 2, Dosing Day, Day 1 Post-Dose, and Weeks 1, 2, 4, and 8 Post-Dose. Self-report surveys and clinician-rated assessments are administered at the study site at Baseline 1, Baseline 2, dosing day, and then post-dose on Day 1 and Weeks 1, 2, 4, and 8. Ambulatory assessments including EMA and voice diary, are administered during the two weeks preceding and the two weeks following dosing, with additional follow-up weekly assessments conducted up to the eighth week of the study. For methodological development purposes, participants are also invited to complete an additional 2 weeks of EMA from Week 2 to Week 4 post-psilocybin. EAR data are collected in three 4-day periods: at Baseline 1, at Week 1 Post-Dose, and Week 4 Post-Dose, with each period comprising 2 weekdays and 2 weekend days.

**Figure 1.**
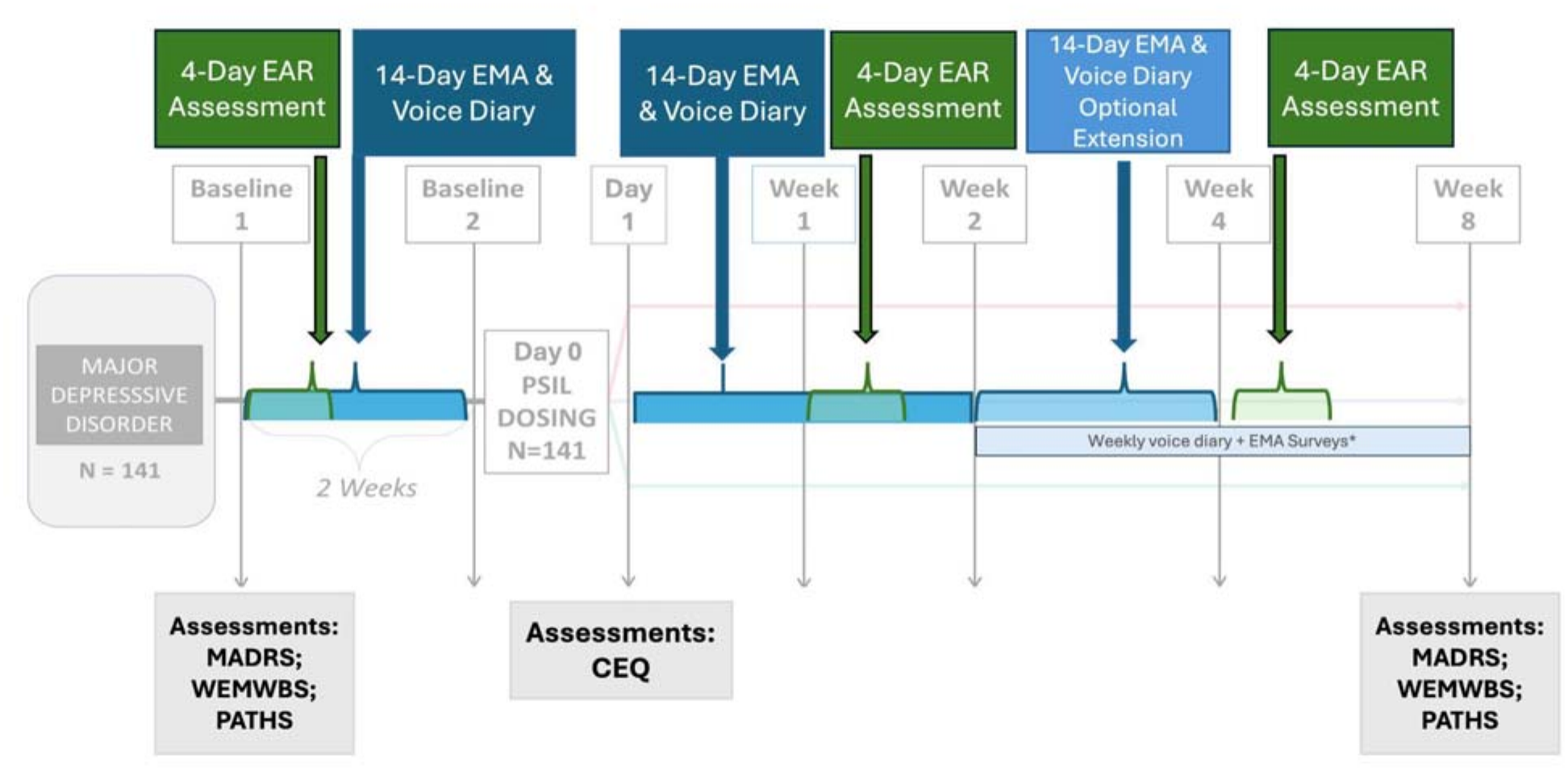
Dial-In Study Timeline and Design. *Note*. EMA = ecological momentary assessment; EAR = Electronically Activated Recorder; CEQ = Challenging Experience Questionnaire; MADRS = Montgomery–Åsberg Depression Rating Scale; WEMWBS = Warwick–Edinburgh Mental Well-Being Scale; PATHS = Psychedelic-Assisted Therapy Harms and Side Effects Scale. EMA and voice-diary assessments are administered four times daily during intensive assessment periods, with the final daily assessment including the voice diary. *An additional, optional 14-day period of EMA and voice diary assessments is offered to participants who complete the first 28 days, extending intensive assessment through Week 4 post-dose. At the end of the daily assessment period, participants answer weekly questions through Week 8 (See Figure 1 and Table S1)

### Ambulatory Assessment Methods

At enrollment, participants are oriented to the ambulatory assessment procedures, including their purpose, frequency, duration, content, privacy and confidentiality protections and compensation, and are given the opportunity to ask questions. Method-specific setup and training are provided before data collection begins, as described below in detail. Because voice-diary and EAR data are not continuously monitored and may not be reviewed until well after collection, participants are informed that these methods cannot be used to relay urgent safety-related information (e.g., harm to self or others) to the research team and are provided with alternative procedures for alerting urgent concerns.

### Ecological Momentary Assessment (EMA) and Voice Diary

Ecological momentary assessment (EMA) involves repeated self-reported assessments of individuals’ current experiences, behaviors, and contexts in real time as they go about their daily lives, minimizing reliance on retrospective recall [15, 49]. In the Dial-In Study, a series of standardized EMA items and voice-diary data are collected using Fabla^1^ [50], a smartphone-based platform for mixed method EMA developed and hosted at Emory University (See Figure 2). On enrollment into the study, participants are informed about the EMA assessment and voice diary, their frequency, content, duration, confidentiality, and compensation schedule. At their baseline visit, participants are instructed to download the Fabla app and to link it to their study ID. Participants are provided with a brief training and orientation to ensure comfort with the app, including a practice survey with the opportunity to ask questions. During this visit, a researcher also works with each participant to develop two personalized EMA items, which are added to the standardized EMA questionnaire, described below.

**Figure 2.**
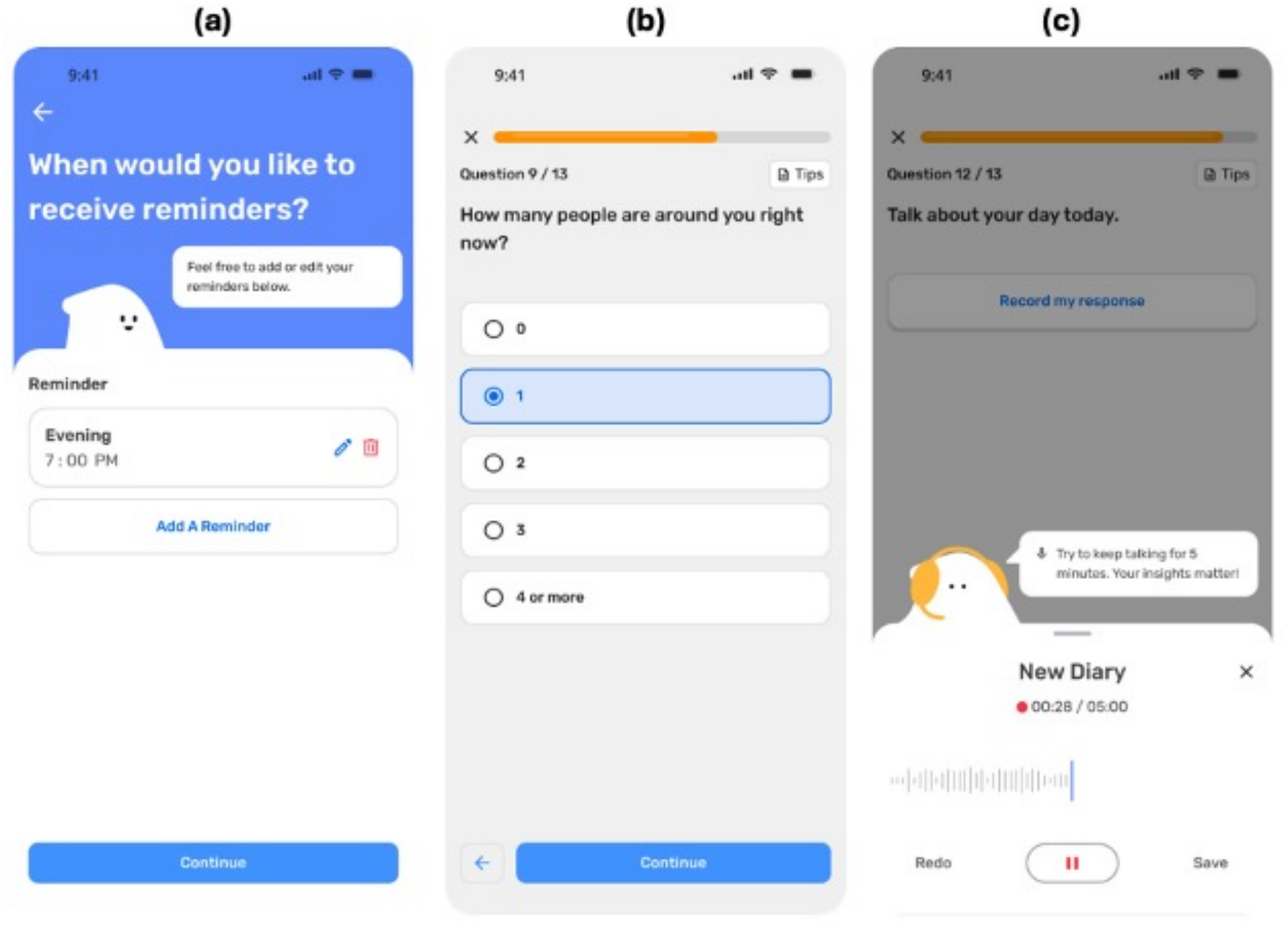
Fabla Smartphone Interface for Ecological Momentary Assessment and Voice Diary Data Collection. ***Note.*** Representative screenshots of the Fabla smartphone platform used for ambulatory data collection in the Dial-In Study. (a) Participants select reminder times for completing scheduled assessments. (b) Participants complete ecological momentary assessment (EMA) items using the smartphone interface. (c) The end-of-day assessment includes a voice diary prompt that allows participants to record an audio response. EMA = ecological momentary assessment.

EMA and voice diaries are administered in intensive bursts aligned with key phases of the trial. EMA and voice-diary assessments begin 14 days prior to dosing, resume following Day 1 post-dose for another 14 days (yielding 28 days total). An additional, optional 14-day period of EMA and voice diary assessments is offered to participants who complete the first 28 days, extending intensive assessment through Week 4 post-dose. At the end of the daily assessment period, participants answer weekly questions through Week 8 (See Figure 1 and Table 1). In addition to compensation for the parent trial, participants are compensated up to $275 for participation related to the Dial-In Study, using a progressive payment schedule that rewards greater adherence and a bonus payment during each EMA/voice diary collection period for >70% adherence.

**Table 1.**
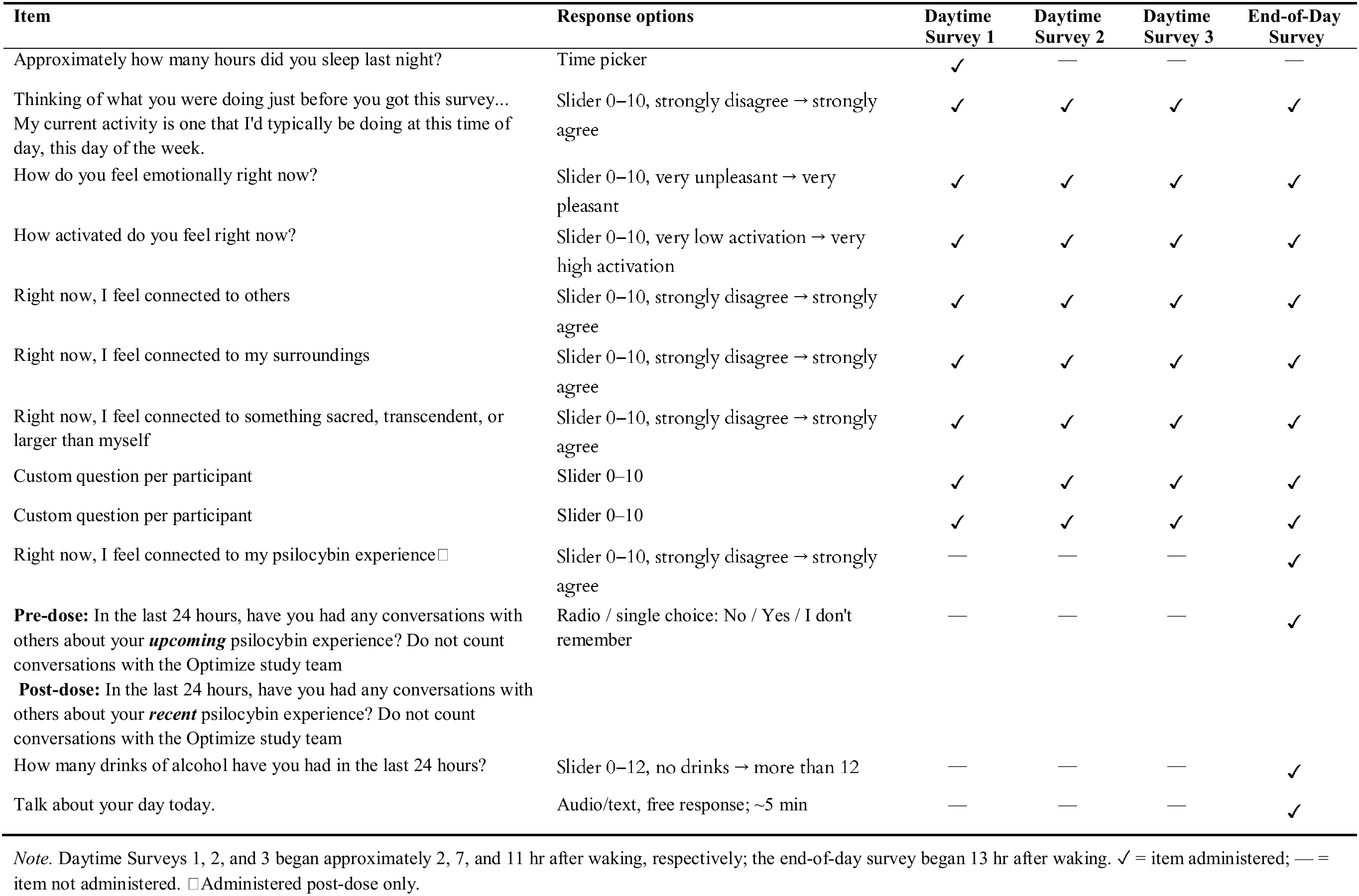
Items and Response Options Across Daily EMA Surveys.

| **Item** | **Response options** | **Daytime Survey 1** | **Daytime Survey 2** | **Daytime Survey 3** | **End-of-Day Survey** |
|---|---|---|---|---|---|
| Approximately how many hours did you sleep last night? | Time picker | ✓ | — | — | — |
| Thinking of what you were doing just before you got this survey...<br>My current activity is one that I'd typically be doing at this time of day, this day of the week. | Slider 0–10, strongly disagree → strongly agree | ✓ | ✓ | ✓ | ✓ |
| How do you feel emotionally right now? | Slider 0–10, very unpleasant → very pleasant | ✓ | ✓ | ✓ | ✓ |
| How activated do you feel right now? | Slider 0–10, very low activation → very high activation | ✓ | ✓ | ✓ | ✓ |
| Right now, I feel connected to others | Slider 0–10, strongly disagree → strongly agree | ✓ | ✓ | ✓ | ✓ |
| Right now, I feel connected to my surroundings | Slider 0–10, strongly disagree → strongly agree | ✓ | ✓ | ✓ | ✓ |
| Right now, I feel connected to something sacred, transcendent, or larger than myself | Slider 0–10, strongly disagree → strongly agree | ✓ | ✓ | ✓ | ✓ |
| Custom question per participant | Slider 0–10 | ✓ | ✓ | ✓ | ✓ |
| Custom question per participant | Slider 0–10 | ✓ | ✓ | ✓ | ✓ |
| Right now, I feel connected to my psilocybin experience□ | Slider 0–10, strongly disagree → strongly agree | — | — | — | ✓ |
| **Pre-dose:** In the last 24 hours, have you had any conversations with others about your ***upcoming*** psilocybin experience? Do not count conversations with the Optimize study team<br>**Post-dose:** In the last 24 hours, have you had any conversations with others about your ***recent*** psilocybin experience? Do not count conversations with the Optimize study team | Radio / single choice: No / Yes / I don't remember | — | — | — | ✓ |
| How many drinks of alcohol have you had in the last 24 hours? | Slider 0–12, no drinks → more than 12 | — | — | — | ✓ |
| Talk about your day today. | Audio/text, free response; ~5 min | — | — | — | ✓ |

*Note.* Daytime Surveys 1, 2, and 3 began approximately 2, 7, and 11 hr after waking, respectively; the end-of-day survey began 13 hr after waking. ✓ = item administered; — = item not administered. □Administered post-dose only.

During intensive assessment periods, participants receive four EMA surveys per day via app notifications, customized to their self-reported sleep schedule, with assessments at approximately 2, 7, 11, and 13 hours after waking. Each survey is open for two hours. The final EMA assessment of each day includes a daily voice diary entry, in which participants are prompted to talk about their experience that day, as though sending a voice memo to a friend, recording for approximately five minutes. Participants who prefer to type rather than speak their daily diary are provided with a text-box response option.

EMA questionnaires are designed to be brief while covering multiple domains relevant to depression and recovery. The standardized item set includes measures of current context and activity; affective valence and activation, and connectedness to others, their surroundings, and something sacred, transcendent, or larger than themselves. See Table 1 for the full list of items. Participants submit their answers on a slider in the application from 0 “strongly disagree”/ “very unpleasant”/ “very low” to 10 “strongly agree”/“very pleasant”/ “very high”. The first assessment of each day additionally queries the number of hours of sleep during the previous night (drop-down using integers). The final assessment of each day queries substance use in the previous 24 hours (checklist), and whether participants discussed their upcoming or completed psilocybin session.

The pre- and post-dose EMA item sets are largely parallel, with several treatment-specific additions following dosing. Post-dose assessments additionally assess participants’ present-moment connection to their psilocybin experience. Two personalized EMA items are created for each participant, enabling idiographic assessment. Participants identify one personally meaningful target that they would like to increase (e.g., joy, motivation, relaxation) and one target that they would like to decrease (e.g., sadness, pain, loneliness). The research team reviews each target to ensure that they are sufficiently specific, likely to vary over the span of the day, readily reportable in the moment, and unlikely to produce discouraging or iatrogenic effects (see supplemental Table S1). The resulting items are incorporated into each participant’s otherwise standardized EMA item set.

The final EMA assessment of each intensive-assessment day also includes a voice-diary component designed to capture participants’ naturalistic reflections on their daily experiences. Participants respond to two open-ended prompts: one asking them to describe their day and a second asking them to reflect on their psilocybin experience prospectively or retrospectively, allowing them to provide a brief stream-of-consciousness audio reflection on their daily experiences. An overview of the daily data collection schedule can be found in Table 1 and Table S2.

Recordings are automatically transcribed using AWS integrated into the Fabla app and subsequently reviewed and corrected by trained research assistants to ensure accuracy. Voice-diary responses are coded using the Verbal Observation of Inner Coping and Experience framework [51], which characterizes affective, cognitive, interpersonal, behavioral, and contextual features of participants’ reports. The individual audio recording serves as the primary unit of coding, and both the transcript and the original audio are used to assign codes to capture linguistic and paralinguistic features of speech that can increase interpretation accuracy. Coding categories capture affective, cognitive, interpersonal, behavioral, and contextual features of participants’ reports. Categories are coded using binary or dimensional response formats, with many dimensional categories rated on a 0–5 scale. The coding system for the VOICE framework is publicly available on OSF (OSF link) and further explained in a tutorial [51]. Recordings are each independently coded by two coders, with regular coder meetings and quality-control checks used to maintain consistent application of the coding framework and minimize coder drift. Category-level interrater reliability is evaluated using Cohen’s weighted kappa, with Cohen’s weighted kappa of 0.41 or higher (corresponding with conventional thresholds for moderate agreement) used to evaluate acceptable agreement.

### Electronically Activated Recorder (EAR)

The Electronically Activated Recorder (EAR) is an ambulatory assessment method that intermittently samples brief segments of ambient sound from participants’ naturalistic environments, providing naturalistic observational data on daily behaviors, social interactions, and environmental context [52–54]. The EAR has been used widely over the last two decades in research on both clinical and nonclinical populations and has generally demonstrated high acceptability and low ratings of obtrusiveness in populations ranging in age from children to older adults [55, 56].

EAR data are collected for three 4-day periods, at Baseline 1 pre-dose, and Week 1 and Week 4 post-dose, with each period comprising two weekdays and two weekend days (see Table S3). During each EAR collection period, participants are given an Android smartphone with the EAR app preloaded and are instructed to wear the device while going about their daily activities. The EAR app records bursts of 30-second samples of ambient audio every 12 minutes silently and without participant awareness of when it is recording. A 6-hour blackout window is also programmed to coincide with each participant’s typical sleep period. This sampling schedule captures a random sample of 5% of the day (approximately 300 audio recordings per four-day period). This recording schedule has been used widely in prior research using this method [52, 53, 57], and found to provide a representative sample of a person’s day with acceptable base rates of key behaviors and features of the wearer’s social environment, while leaving 95% of the participants’ daily life unrecorded and private.

Participants are provided instructions for device use (e.g., not covering the device with clothing, allowing the “recording” symbol to be visible) and privacy, including use of a privacy button that prevents recording for 30 minutes and the option to leave behind the device during activities they do not wish to have recorded. Participants are also given an opportunity to review and delete recordings before returning the device. EAR data are stored locally on the smartphone and then downloaded to a secure server when the EAR devices are returned to the research team. EAR recordings are coded by trained human coders using a study-specific adaptation of the *Social Environment Coding of Sound Inventory* (SECSI) [58], a behavioral coding framework designed to characterize acoustically detectable features of participants’ everyday environments, activities, behaviors, and social interactions. Coding domains include physical and social context, interaction partners, conversation topics and types, daily activities, emotional expression and affect, and interpersonal behaviors. Categories characterize, among other features, whether participants are alone or with others, whom they are interacting with, whether conversations are practical or substantive, and whether interactions involve conflict or tension. Additional categories capture participants’ emotional and interpersonal behavior, including affection and gratitude, self-criticism, frustration, support seeking, support giving, and support receiving, as well as study-specific questions. Behaviors expressed by others in participants’ immediate environments are also coded, including affection, humor, praise and encouragement, validation, sharing of feelings, dismissiveness or minimizing, criticism, hostility, and emotional overinvolvement. All categories are binarily coded (0 = the feature is not present in the audio file, 1 = the given feature is present). For more information about the EAR Coding System, see this OSF page. Participant speech captured in EAR recordings is additionally transcribed for linguistic analysis. Transcribed speech is analyzed using Linguistic Inquiry and Word Count (LIWC-22) [59] to derive count-based linguistic indicators, including self- and other-focused language.

## Measures

### Self-Report and Clinician Rated-Measures

#### Warwick-Edinburgh Mental Well-Being Scale (WEMWBS)

The WEMWBS is a 14-item self-report measure of psychological well-being that captures both emotional and functional aspects of well-being [60]. Example items include “I’ve been feeling optimistic about the future,” “I’ve been dealing with problems well,” and “I’ve been feeling close to other people.” Items are positively worded and rated on a 5-point scale ranging from 1 (*none of the time*) to 5 (*all of the time*), with participants reporting on their experiences over the previous 2 weeks. Higher scores indicate greater mental well-being. The WEMWBS is administered pre-dosing at Baseline 1, Baseline 2, and post-dosing at Weeks 1, 2, 4, and 8 as part of the OPTIMIZE parent study procedure. For our hypothesis testing, we will use participants’ WEMWBS scores at Baseline 1 and Week 8 post-dosing.

#### Montgomery–Åsberg Depression Rating Scale (MADRS)

The MADRS is a 10-item clinician-rated measure of depressive symptom severity designed to be sensitive to changes in symptoms over the course of treatment [61]. The structured interview [62] assesses reported and apparent sadness, inner tension, reduced sleep and appetite, concentration difficulties, lassitude, inability to feel, pessimistic thoughts, and suicidal thoughts. Each item is rated from 0 to 6, yielding a total score ranging from 0 to 60, with higher scores indicating greater depressive symptom severity. MADRS assessments are administered by trained independent raters who are blinded to treatment allocation and visit number. For our hypothesis testing, we will use participants’ MADRS scores at Baseline 1 and Week 8 post-dosing.

#### Challenging Experience Questionnaire (CEQ)

The CEQ is a 26-item self-report measure designed to assess challenging experiences associated with psilocybin use [63]. The measure comprises seven dimensions of challenging experience: fear, grief, insanity, death, isolation, physiological distress, and paranoia. Example items assess experiences such as fear, feelings of isolation, feeling as though one is dying, and concern about losing one’s sanity. The CEQ is administered at Day 1 and Week 8 post-dose as part of the OPTIMIZE parent study procedure. For this study, we will use the Day 1 CEQ answers in the analyses.

#### Psychedelic-Assisted Therapy Harms and Side Effects Scale (PATHS)

The PATHS assesses potential adverse effects that may occur in psychedelic-assisted therapy [64]. The measure captures up to 54 discrete adverse effect criteria. Participants are instructed to rate the extent to which they are experiencing each potential adverse effect on a scale from 0 (“None/Does not apply”) to 3 (“Quite a bit”). An example is “I have been struggling with the meaning or purpose of my life”. Items predicated on experiences specific to psychedelic treatment, such as inappropriate physical contact during dosing, are omitted from the baseline measure. For our hypothesis testing, we will use participants’ PATHS scores at Baseline 1 and Week 8 post-dosing.

### Analysis Plan

Analyses will be conducted using R 4.6. [65]. The analytic sample will include participants who complete pre-dose study activities, receive a psilocybin dose, and provide at least one post-dose assessment of the relevant outcome. Missing EMA time-series observations will be imputed using the *imputeTS* R package [66, 67]. Missing data on repeatedly assessed continuous outcome variables will be handled using full-information maximum likelihood under the assumption that data are missing at random. Variables will be transformed as appropriate when distributional violations of model assumptions can be addressed through transformation, with all transformations reported transparently. Participants contributing fewer than 50 codable EAR sound files at each given timepoint will be excluded from analyses involving EAR-derived variables. Parent-trial treatment conditions will be included as a covariate in planned analyses. Demographic and baseline characteristics will be summarized using descriptive statistics. Statistical tests will use a two-sided significance level of .05, and effect estimates will be reported with 95% confidence intervals.

### Power Analysis

Statistical power was evaluated for the primary linear mixed-effects model testing the cross-level interaction between time (baseline and week 8) and a binary variable (e.g., sudden gain). Assuming a total sample size of *N* = 141 individuals, a standard moderate effect size (Cohen’s f2 = 0.06) for the interaction, a two-sided significance level of α = 0.005 (adjusting for 9 multiple tests in the most restrictive case), and a standard moderate-to-high baseline-to-follow-up correlation of r = 0.50 typical of longitudinal behavioral data, the study achieves approximately 99% statistical power to detect the interaction effect.

### Outcome Construction

Across hypotheses, outcomes will be characterized using depressive symptoms, psychological well-being, and psychedelic-related adverse effects. MADRS and WEMWBS scores will characterize depressive-symptom and well-being outcomes, respectively, whereas the sum of worsening domains on the PATHS from pre-to post-dosing will characterize adverse-effect outcomes.

### Construction of Risk and Protective Variables

#### EMA-Derived Sudden Gains and Losses

We will derive sudden gains and losses from each participant’s EMA time series, using momentary affective valence as the primary state variable. The ts_levels() function in the *casnet* R package will identify abrupt shifts within each individual trajectory. We will specify a minimum data split of 12 observations, corresponding to approximately 3 days of EMA assessments; a minimum level duration of 28 observations, corresponding to approximately 1 week; and a change-sensitivity parameter of .05. We will classify upward abrupt shifts as sudden gains and downward abrupt shifts as sudden losses.

#### EAR-Derived Risk and Protective Factors

We will aggregate dichotomous EAR behavioral codes to create continuous indicators of participants’ everyday behaviors and social environments (i.e., for each timepoint, the percentage of the four-day period during which each feature of behavior and social environment occurred). Linguistic analyses of transcribed EAR speech will provide complementary indicators of self- and other-focused language.

The EAR-derived variables used in H2 will capture social support, interpersonal conflict, and conversational focus on the self-versus others. Social support will serve as a protective factor and interpersonal conflict as a risk factor. We will operationalize conversational focus using self- and other-focused language, with greater movement from self-focused toward other-focused language representing the hypothesized protective pattern.

#### Voice Diary Derived Risk and Protective Factors

The voice-diary analyses will focus on personal reflection/perspectival flexibility and rigidity, indexed through thematic coding using the VOICE as well as linguistic analyses of speech features contained in the transcripts. Indicators of curiosity, perspective-taking, and explorative language will contribute to the personal reflection/perspectival flexibility exploration construct, whereas ruminative and repetitive language will index rigidity. For an overview of the construction of risk and protective variables, please see Table 2.

**Table 2.** Construction of Risk and Protective Variables.

| Variable | Role | Data source | Construction and operationalization |
|---|---|---|---|
| Sudden gains | Protective | EMA | Upward abrupt shifts in participants’ momentary affective valence time series. Affective valence is assessed four times daily and serves as the primary state variable for detecting transitions. Abrupt shifts are identified at the individual level using the ts_levels() function in the casnet R package. |
| Sudden losses | Risk | EMA | Downward abrupt shifts in participants’ momentary affective valence time series, identified at the individual level using the same ts_levels() procedure used to identify sudden gains. |
| Social support | Protective | EAR | Behavioral indicators of social support coded from EAR recordings using the Social Environment Coding of Sound Inventory (SECSI). Dichotomous snippet-level behavioral codes are aggregated into continuous variables expressed as proportions of total coded snippets. |
| Conflict | Risk | EAR | Behavioral indicators of interpersonal conflict coded from EAR recordings using the SECSI. Dichotomous snippet-level codes are aggregated into continuous variables expressed as proportions of total coded snippets, with greater conflict hypothesized to predict poorer outcomes. |
| Shift in conversational focus from self to others | Protective | EAR | Derived from transcribed EAR recordings using Linguistic Inquiry and Word Count (LIWC-22) indices of self- and other-focused language. Greater relative conversational focus on others is hypothesized to predict better outcomes. |
| Personal reflection / perspectival flexibility | Protective | Voice diary | Derived from nightly voice-diary responses to prompts about participants’ daily experiences and their psilocybin experience. Curiosity, perspective-taking, and explorative language are used as indicators of personal reflection and perception-box exploration. |
| Rigidity | Risk | Voice diary | Derived from nightly voice-diary responses, with ruminative and repetitive language used as indicators of rigidity. Greater rigidity is hypothesized to predict poorer outcomes. |

*Note.* EMA = ecological momentary assessment; EAR = Electronically Activated Recorder; SECSI = Social Environment Coding of Sound Inventory; LIWC-22 = Linguistic Inquiry and Word Count 2022 [59]. Risk and protective classifications reflect the hypothesized direction of association with post-psilocybin outcomes. Sudden gains and losses are derived primarily from momentary affective valence; additional EMA variables may be examined in supplementary or exploratory dynamic-systems analyses.

### Hypothesis Testing

For H1, participant-level EMA time series will be analyzed using dynamic-systems methods implemented in the *casnet* R package. Sudden gains or losses will be estimated as binary variables indicating presence or absence. Across 6 mixed-effects models, one of the three endpoints (i.e., depressive symptoms, psychological well-being, and psychedelic-related adverse effects) will serve as the outcome variable, and predictors include a time variable (baseline vs. week 8) nested within individuals, one of the abrupt shifts (i.e., sudden gain or sudden loss), lag of the abrupt shift (time between the occurrence of shift and the week 8 assessment), interaction of the abrupt shift and time, and other covariates. The interaction regression coefficients – abrupt shifts X time – will constitute the primary tests of whether within-person dynamic changes in EMA trajectories are associated with change in outcomes.

For H2a, risk (interpersonal conflict) and protective factors (social support, conversation focus) measured at three time points (Baseline, Week 1, and Week 4) will be examined simultaneously as longitudinal predictors of depressive symptoms, psychological well-being, and psychedelic-related adverse effects in 9 separate mixed-effects models. Each model will have one of the endpoints as an outcome, and predictors include a time variable (baseline vs. week 8) nested within individuals, an EAR predictor assessed at three times, interaction term of an EAR predictor and time, and other covariates. The interaction regression coefficients – EAR predictor X time – will constitute the primary tests of whether risk and protective factors are associated with change in outcomes.

For H2b, voice diary variables (rigidity, personal reflection/perspectival flexibility) aggregated at two time points (before dosing vs. after dosing) will be examined simultaneously as longitudinal predictors of depressive symptoms, psychological well-being, and psychedelic-related adverse effects in 6 separate mixed-effects models. The procedures will be similar to above.

For H3a, three linear regression models will examine associations between challenging experiences during psilocybin dosing, operationalized using CEQ scores assessed at Day 1 post-dosing, and week-8 depressive symptom, psychological well-being, and psychedelic-related adverse-effect outcomes, controlling for baseline values. For H3b, moderation effects will be tested by including interaction terms, one at a time, between CEQ scores and the previously tested risk and protective factors, including sudden gains and losses and factors derived from EAR, EMA, and voice-diary data.

To account for multiple testing, we will control the false discovery rate (FDR) using the Benjamini-Hochberg procedure within families of conceptually related hypothesis tests (i.e., EAR, EMA) [68]. Families of tests will be defined a priori based on the hypothesis being tested and the source of the predictor variable. In that vein, the six tests of EMA-derived sudden gains and losses corresponding to H1 will constitute one family; the nine tests of EAR-derived risk and protective factors in H2a will constitute a second family; the six tests of voice-diary derived factors in H2b will constitute a third family. For H3a, the three tests of the association between CEQ and outcomes will constitute a fourth family; for H3b, for each moderator, the three tests of moderation effects (one for each outcome) will constitute a separate family.

Exploratory analyses will examine time-varying effects of voice diary variables on each of the three outcomes by fitting models of change over time. For other exploratory aims, additional dynamic-systems indices will be examined as moderators of associations between risk and resilience variables derived from EMA, voice-diary, and EAR data and clinical outcomes. Additional analyses will examine associations between the timing of dynamic changes relative to psilocybin dosing and subsequent clinical outcomes. Exploratory analyses will also examine additional EMA variables, including affective activation, activity prototypicality, social connectedness, and participant-specific items, as well as acoustic voice biomarkers derived from voice-diary and EAR recordings.

### Patient and Public Involvement

Patients and members of the public were not involved in the design, conduct, reporting or dissemination plans of this study.

### Ethics and Dissemination

The project was approved by WCG Institutional Review Board, with Emory University relying on WCG as the reviewing board. All participants provide written informed consent. Findings will be disseminated through peer-reviewed publications and scientific presentations.

## Discussion

The Dial-In Study is designed to characterize predictors of differential response to psilocybin, including both improvement and worsening following treatment. Its primary contribution is the use of multi-method, ambulatory, intensive longitudinal assessment to examine changes that unfold around psilocybin dosing and within participants’ everyday lives. By combining EMA, voice-diary, EAR, and repeated assessments of depression, well-being, and adverse effects, the study will examine subjective, behavioral, interpersonal, and clinical variables across different time scales rather than relying only on assessments conducted at discrete study visits.

This design makes it possible to examine post-acute change at a finer temporal level. Intensive longitudinal assessment allows sudden gains and losses to be identified within individual affective trajectories, extending work showing that improvement and worsening in psychotherapy can occur discontinuously rather than gradually [38, 39]. Ambulatory assessments administered in the contexts of participants’ daily lives also allow these changes to be considered alongside participants’ everyday environments and behaviors. EAR data characterize social support, interpersonal conflict, and conversational focus, while voice-diary data capture personal reflection and rigidity. The inclusion of personalized EMA items further allows participants’ own desired outcomes to be represented alongside standardized measures. Together, these methods allow the study to examine processes that would be difficult to capture through conventional baseline and follow-up assessments alone.

The resulting data may help clarify several sources of differential response to psilocybin. First, they will allow examination of whether sudden gains and losses precede differences in depression, well-being, and adverse effects. Second, they will allow risk and protective factors observed in participants’ daily lives to be examined as predictors of these same outcomes. Finally, Dial-In may help clarify why challenging dosing experiences are followed by improvement in some individuals and worsening in others. Challenging experiences have been associated with poorer outcomes, but these associations are inconsistent, and difficult experiences can also be followed by perceived benefit [19, 20, 25]. By examining whether risk and protective factors occurring before and after dosing moderate this relationship, the study may provide a more detailed account of the conditions under which challenging experiences are followed by better versus worse outcomes.

Several strengths and limitations should be considered. The prospective design and repeated assessment surrounding dosing allow temporal ordering of predictors and subsequent outcomes, while the combination of self-report, naturalistic speech, and observed everyday behavior provides complementary information about participants’ post-acute experiences. At the same time, Dial-In is observational, and associations between risk and protective factors and later outcomes cannot establish causality. Intensive EMA, voice-diary, and EAR assessment may also result in missingness or differential adherence, and the different assessment schedules of these methods limit the extent to which all data streams can be aligned at the same temporal scale. In addition, the study is embedded within a clinical trial of adults with major depressive disorder, which may limit generalizability beyond this treatment context.

More broadly, Dial-In will generate fine-grained longitudinal data that may support continued investigation of temporal and dynamic predictors of differential response to psilocybin. Identifying patterns that consistently precede improvement or worsening could inform future research on early indicators of vulnerability and resilience and on the periods in which additional support may be most relevant. The present study will not establish how such factors should be modified in treatment, but it may provide a basis for more targeted research on how post-acute experiences, behaviors, and environments contribute to outcomes following psilocybin.

## Author Contributions

**Andreea Sutu:** Writing – original draft; review and editing; Methodology, Investigation, Project administration, Supervision, and Visualization.

Led the drafting and development of the manuscript, contributed to the refinement of the study methodology and analytical framework, supported study implementation and data collection, coordinated research activities and manuscript preparation, supervised research personnel, and developed manuscript figures and visualizations.

**Deanna Kaplan:** Conceptualization; Methodology, Supervision, Funding acquisition, Investigation, Project Administration, Resources Writing – review & editing.

Contributed to the original conceptualization and methodological design and implementation of the Dial-In study; provided scientific oversight and supervision of research activities; supported data collection and project coordination; contributed to securing funding and research resources; critically reviewed and revised the manuscript.

**Christina Sauder:** Funding acquisition, Investigation, Project administration, Resources, Writing – review & editing.

Contributed to securing research funding and resources, supported study implementation and coordination, participated in research and data collection activities, and critically reviewed and revised the manuscript. Contributed to research administration and operational coordination at the Vail Health site, facilitating study implementation, research logistics, and the coordination of personnel and resources necessary for study conduct.

**Charles L. Raison:** Funding acquisition, Investigation, Project administration, Resources, Supervision, Writing – review & editing.

Served as Principal Investigator of the parent OPTIMIZE clinical trial, providing scientific and clinical oversight of study design, implementation, and regulatory compliance; contributed to securing funding and research resources, supported study conduct and coordination, and critically reviewed and revised the manuscript.

**Santiago Jose Arconada Alvarez:** Software; Methodology; Investigation; Writing - review and editing.

Developed the Fabla software platform, contributed to its adaptation and implementation for the Dial-In study, and collaborated on the development and execution of data collection procedures.

**Zainab Salako:** Investigation; Project administration, Writing - review and editing

Contributed to participant coordination and study implementation at the Emory site, supported data collection activities, and trained research coders involved in behavioral data coding.

**Nicole Pozzo:** Methodology; Investigation; Project administration; Supervision; Study design.

Contributed to the development and refinement of the EAR and VOICE behavioral coding schemes, supported participant coordination and data collection at the Emory site, and trained and supervised research coders involved in behavioral data coding.

**Zhuo Job Chen:** Methodology; Data Analysis Planning, Writing - Review & editing.

Developed the statistical analysis strategy and analytical framework for the Dial-In study, including approaches to evaluating longitudinal data and clinical outcomes, and contributed to the critical review and revision of the manuscript.

**Roman Palitsky:** Conceptualization; Methodology; Supervision; Funding acquisition; Writing – original draft; review & editing.

Contributed to the original conceptualization and methodological design of the Dial-In study, provided scientific oversight and supervision, contributed to securing research funding, supported manuscript development and drafting, and critically reviewed and revised the manuscript.

## Data Availability Statement

Deidentified data, subsets of raw data and statistical analysis scripts will be available upon request.

## Footnotes

[1] Fabla is a HIPAA-compliant smartphone application allowing real-time customization and study-specific tailoring without reliance on third-party platforms.

