## Supplemental Material for "How Does Psilocybin Affect Daily Life Experience and Behavior? Protocol For a Multimethod Ambulatory Assessment Study Within a Clinical Trial of Psilocybin for Depression"

**Supplemental A**

**Parent Study Design & Procedure**

The OPTIMIZE Study (trial number NCT06512194) is a Phase 2a study examining whether post-psilocybin transcutaneous auricular vagus nerve stimulation (taVNS) enhances and extends the therapeutic effects of psilocybin in major depressive disorder. The study will enroll up to 141 adults aged 18–70 years with a current major depressive episode of at least 60 days’ duration and moderate or greater severity (MADRS ≥28), with the goal of obtaining approximately 120 evaluable participants. All participants receive a single open-label 25-mg dose of psilocybin within a standardized set-and-setting framework that includes preparatory sessions, therapist support during dosing, and post-session integration therapy.

Prior to dosing, participants complete two preparation sessions with study facilitators, each lasting up to two hours. Preparation includes discussion of participants’ life histories, personally relevant themes, expectations, and logistical aspects of the dosing session. On dosing day, participants ingest 25 mg of psilocybin after completion of pre-dose safety procedures and are monitored for a minimum of eight hours by two trained facilitators. Following dosing, participants complete three supportive, non-directive integration sessions occurring approximately 1 day, 1 week, and 2 weeks post-dose. These sessions provide opportunities to reflect on the dosing experience and integrate personally salient insights into everyday life.

Following dosing, participants are randomized (1:1:1) to receive seven days of twice-daily taVNS, sham taVNS, or treatment-as-usual consisting of integration sessions alone. Participants assigned to active or sham taVNS complete twice-daily, approximately 20-minute at-home sessions for seven consecutive days beginning after the Day 1 integration session. Stimulation sessions are paired with prompts to recall personally salient aspects of the psilocybin experience and music from the dosing-session playlist. The first two sessions are monitored by study personnel, with daily safety check-ins for the remainder of the seven-day period. Participants assigned to no taVNS complete the same post-dose integration and follow-up schedule without stimulation.

Study assessments are collected at screening, Baseline 1, Baseline 2, and at 1 day and 1, 2, 4, and 8 weeks post-dosing, as indicated in Figure 1. Clinical and psychological outcomes including depression, anxiety, well-being, functioning, quality of life, psychological flexibility, connectedness (with others and with the world), suicidal ideation, and adverse events are assessed from baseline through 8 weeks post-dosing. Self-reported salience of memory for the psychedelic experience will be assessed at Day 2, Week 1, and Week 8. Perceived impact of the psychedelic experience on measures related to wellbeing is assessed at post-psilocybin dosing Weeks 1 and 8. The primary endpoint is change in Montgomery–Åsberg Depression Rating Scale (MADRS) scores from baseline to 8 weeks.

Participants are asked to comply with study-related behavioral restrictions designed to support participant safety and reduce confounding influences on dosing outcomes. These include temporary abstinence from alcohol, tobacco, and illicit psychoactive substances surrounding dosing, and medication restrictions as determined by study investigators. Reproductive precautions are required where applicable.

**Supplemental B**

**Table S1**

*Guidelines for Developing Personalized Ecological Momentary Assessment Items*

| Item-creation criterion | Guidance |
| --- | --- |
| Collaborative development | Two personalized EMA items are developed collaboratively by the participant and study clinician to reflect the participant’s personal goals for psilocybin treatment. |
| Response scale | Each item is rated on a 0–10 scale, with 0 representing the lowest possible level of the target construct and 10 representing the highest possible level. |
| Response anchors | The meaning of scores of 0 and 10 is defined explicitly for each personalized item to support consistent interpretation across repeated assessments. |
| Positive goal | One item reflects a strength or positive outcome that the participant hopes to increase (e.g., well-being, motivation, focus). |
| Challenge or concern | One item reflects a challenge or concern that the participant hopes to decrease or manage (e.g., anxiety, stress, negative thoughts). |
| Suitability for intensive assessment | Targets should be meaningful and appropriate for assessment multiple times per day. Items should assess experiences that may vary within a day rather than relatively fixed or once-daily experiences, such as the previous night’s sleep. |
| Documentation and submission | The finalized item wording and response anchors are documented and submitted to the EMA designer within 3 days of the Baseline Visit. |
| Example: positive goal | Goal: Increase motivation throughout the day. Item: “How motivated do you feel to engage with activities right now?” 0: “Not at all motivated, no interest in doing anything.” 10: “Extremely motivated, eager to do everything planned for the day.” |
| Example: challenge/concern | Goal: Reduce anxiety or worry. Item: “How anxious or worried do you feel at this moment?” 0: “No anxiety, calm and relaxed.” 10: “Extreme anxiety, feeling overwhelmed and unable to relax.” |

*Note.* EMA = Ecological Momentary Assessment. Personalized items are intended to capture participant-specific treatment goals while remaining suitable for repeated momentary assessment.

**Table S2**

*Dial-In EMA Assessment Schedules and Compensation*

| **EMA schedule** | **Pre-dose** | **Weeks 1–2 post-dose** | **Weeks 3–4 post-dose** | **Weeks 5–8 post-dose** | **Max. compensation** |
| --- | --- | --- | --- | --- | --- |
| **Standard** | Daily × 14 days | Daily × 14 days | Weekly | Weekly | $175 |
| **Extended** | Daily × 14 days | Daily × 14 days | Daily × 14 days | Weekly | $275 |

*Note.* Daily assessments are delivered four times per day, with the final daily survey containing the voice diary. Participants who do not complete the optional 14-day extension begin weekly assessments after Week 2; participants who complete the extension begin weekly assessments after Week 4. Maximum compensation assumes completion of all eligible Fabla assessments and receipt of the >70% adherence bonuses.

**Table S3**

*Dial-In EAR Assessment Schedule*

| **EAR assessment period** | **Timing** | **Duration** | **Composition** |
| --- | --- | --- | --- |
| Baseline | Baseline 1 | 4 consecutive days | 2 weekdays + 2 weekend days |
| Post-dose 1 | Week 1 post-dose | 4 consecutive days | 2 weekdays + 2 weekend days |
| Post-dose 2 | Week 4 post-dose | 4 consecutive days | 2 weekdays + 2 weekend days |

*Note.* Each 4-day EAR assessment period comprises two weekdays and two weekend days. During each period, the EAR records 30-second ambient audio samples every 12 minutes, with a 6-hour recording blackout window aligned with the participant’s typical sleep period.
